# A meta-analysis of polygenic risk scores for mood disorders, neuroticism, and schizophrenia in antidepressant response

**DOI:** 10.1101/2021.05.28.21257812

**Authors:** Giuseppe Fanelli, Katharina Domschke, Alessandra Minelli, Massimo Gennarelli, Paolo Martini, Marco Bortolomasi, Eduard Maron, Alessio Squassina, Siegfried Kasper, Joseph Zohar, Daniel Souery, Stuart Montgomery, Diego Albani, Gianluigi Forloni, Panagiotis Ferentinos, Dan Rujescu, Julien Mendlewicz, Diana De Ronchi, European College of Neuropsychopharmacology (ECNP) Pharmacogenomics & Transcriptomics Thematic Working Group, Bernhard T Baune, Alessandro Serretti, Chiara Fabbri

**Author notes:** Corresponding author: Alessandro Serretti, MD, PhD, Department of Biomedical and Neuromotor Sciences, University of Bologna, Viale Carlo Pepoli 5, 40123 Bologna, Italy.

## Abstract

About two-thirds of patients with major depressive disorder (MDD) fail to achieve symptom remission after the initial antidepressant treatment. Despite a role of genetic factors was proven, the specific underpinnings are not fully understood yet. Polygenic risk scores (PRSs), which summarise the additive effect of multiple risk variants across the genome, might provide insights into the underlying genetics. This study aims to investigate the possible association of PRSs for bipolar disorder, MDD, neuroticism, and schizophrenia (SCZ) with antidepressant non-response or non-remission in patients with MDD. PRSs were calculated at eight genome-wide P-thresholds based on publicly available summary statistics of the largest genome-wide association studies. Logistic regressions were performed between PRSs and non-response or non-remission in six European clinical samples, adjusting for age, sex, baseline symptom severity, recruitment sites, and population stratification. Results were meta-analysed across samples, including up to 3,637 individuals. Bonferroni correction was applied. In the meta-analysis, no result was significant after Bonferroni correction. The top result was found for MDD-PRS and non-remission (p=0.004), with patients in the highest vs. lowest PRS quintile being more likely not to achieve remission (OR=1.5, 95% CI=1.11-1.98, p=0.007). Nominal associations were also found between MDD-PRS and non-response (p=0.013), as well as between SCZ-PRS and non-remission (p=0.035). Although PRSs are still not able to predict non-response or non-remission, our results are in line with previous works; methodological improvements in PRSs calculation may improve their predictive performance and have a meaningful role in precision psychiatry.

## 1. Introduction

Major depressive disorder (MDD) is a common psychiatric condition that is among the leading causes of disability worldwide, accounting for a 61.1% increase in the number of disability-adjusted life years (DALYs) over the past two decades (Diseases and Injuries, 2020). Up to 60% of patients with MDD do not respond adequately to the first prescribed antidepressant, requiring either a dose increase, switching to another antidepressant, or augmentation with a different pharmacological agent (De Carlo et al., 2016). Remission is achieved by only 37.5% of patients after six weeks of treatment with first-line antidepressants, and non-responding patients undergoing several consecutive treatment steps achieve lower remission and higher relapse rates (Rush et al., 2006). Therefore, early identification of each patient’s most appropriate treatment might help to reduce the burden of the disease and the related cost to society.

Several socio-demographic and clinical predictors of non-response or non-remission have been identified, including older age, longer duration of the depressive episode, greater severity at baseline, and the presence of anxiety symptoms (Kautzky et al., 2018). Genetic variability may also contribute, as indicated by a single-nucleotide polymorphism (SNP)-based heritability of 13.2% for remission, though the specific loci involved were not identified (Pain et al., 2020).

Polygenic risk scores (PRSs) summarise the additive effect of common genetic risk variants across the genome, and they have shown promising clinical utility in other fields of medicine (Natarajan et al., 2017). Interestingly, people with a PRS for coronary artery disease above the 80^th^ percentile were shown to benefit most from statin treatment in terms of preventing acute cardiac events, with a relative risk reduction of 44% compared to 26% observed in patients with a lower PRS (Natarajan et al., 2017). Using the same approach, we found that higher PRSs for schizophrenia (SCZ) in patients with MDD may be associated with worse response to the first antidepressant treatment, and that individuals having a lower SCZ-PRS showed higher chances of response when antidepressants were not augmented with antipsychotics (Fanelli et al., 2021). A positive genetic correlation was also identified between non-response to antidepressants and neuroticism (NEU), schizotypy, and mood disorders, suggesting the existence of underlying shared genetics (Wigmore et al., 2020).

In light of these findings, we aimed to extend our previous results to other samples (Fanelli et al., 2021), through a large meta-analysis of relevant PRSs (MDD, bipolar disorder (BP), SCZ, NEU) and antidepressant non-response and non-remission in MDD. PRSs could indeed help to better stratify patients with respect to their chances of response/remission and lead to the early implementation of second-line treatment strategies.

## 2. Methods

### 2.1. Target samples

#### 2.1.1. Brescia

This sample included a total of 501 subjects with MDD (Diagnostic and Statistical Manual of Mental Disorders-IV (DSM-IV) criteria) who had been referred to the “Villa Santa Chiara” Psychiatric Hospital in Verona, Italy. Diagnosis of unipolar depression was confirmed using the Structured Clinical Interview for DSM-IV Axis I Disorders (SCID-I). Patients were excluded if they had met criteria for another primary neuropsychiatric disorder or comorbid eating disorder, substance/alcohol abuse or dependency. Treatment non-response was defined as the failure to respond to at least one adequate trial of antidepressants. Genotyping was performed using the Infinium PsychArray-24 BeadChip or the Infinium Multi-Ethnic Genotyping Array (N=215 and 286, respectively). Additional information is available elsewhere (Minelli et al., 2015).

#### 2.1.2. European Group for the Study of Resistant Depression (GSRD)

The sample included 1,346 genotyped patients (Infinium PsychArray-24 BeadChip) with MDD recruited from the European Group for the Study of Resistant Depression (GSRD) as part of a multicentric study. MDD was diagnosed using the Mini International Neuropsychiatric Interview (MINI). Patients were excluded if they had met criteria for another primary psychiatric disorder in the six months prior to enrolment. Treatment response/remission were determined using the Montgomery-Åsberg Depression Rating Scale (MADRS) (50% improvement and MADRS ≤10, respectively). Further details are available elsewhere (Souery et al., 2007). This sample was previously included in a similar PRS study focused on non-response to the last antidepressant and treatment-resistant depression (TRD) (Fanelli et al., 2021).

#### 2.1.3. Münster

It is a naturalistic study of 621 participants aged 18 – 85 years with MDD, as assessed by the SCID-I. Participants were recruited at the Department of Psychiatry, University of Münster, Germany (Baune et al., 2010). Patients with SCZ spectrum disorders, BD, current alcohol or drug dependence, neurological or neurodegenerative illnesses were excluded. Response and remission at week six were measured using the 21-item Hamilton Depression Rating Scale (HAMD_21_) (50% improvement and HAMD_21_ ≤7, respectively). Genotyping was performed using the Infinium PsychArray-24 BeadChip.

#### 2.1.4. Sequenced Treatment Alternatives to Relieve Depression (STAR^*^D)

The STAR*D study was conducted to compare tolerability and efficacy of antidepressants throughout four sequential treatment levels in patients with MDD of at least moderate severity. Symptom severity was assessed using the Quick Inventory of Depressive Symptomatology Clinician-rated scale (QIDS-C_16_) every two weeks; level 1 exit data were considered for this study. Response/remission were defined as a 50% decrease in symptom severity and QIDS-C_16_ ≤5 at week 12, respectively. A total of 1,948 participants were genotyped (Affymetrix GeneChip Human Mapping 500K Array Set or Affymetrix Genome-Wide Human SNP Array 5.0). The study is described in depth elsewhere (Howland, 2008).

#### 2.1.5. Tartu

This sample included 83 outpatients with MDD recruited at the Psychiatric Clinic of the University Hospital of Tartu, Estonia. The diagnosis was made using the MINI 5.0.0, psychiatric history, and medical records. Patients with other primary neurological or psychiatric disorders were excluded from the study. Treatment response/remission were measured using the MADRS, in line with the previous samples. Further information is available elsewhere (Aluoja et al., 2018). The samples sequenced were genotyped using the Illumina 370CNV array. Further information is available elsewhere (Tammiste et al., 2013).

### 2.2. Quality control of the target datasets

Quality control (QC) and population principal component analysis (PCA) were performed through the Ricopili pipeline in each of the six target samples separately (Lam et al., 2020). Single-nucleotide polymorphisms (SNPs) were retained if they had a call rate ≥0.95, differences in call rates between cases and controls (missing difference) ≤0.02, minor allele frequency (MAF) ≥0.01, and Hardy-Weinberg equilibrium p-value ≥1e-6. Individuals were retained if they had an autosomal heterozygosity deviation within ±0.2, call rate ≥0.98, and no genetic/pedigree sex mismatch.

To assess between-subjects relatedness and population stratification, all pairs of individuals with identity-by-descent proportion >0.2 were identified using linkage disequilibrium-pruned data (r^2^ <0.2), and one individual from each pair was removed. PCA was used to determine population stratification (Eigenstrat); population outliers were removed according to the mean ±6 standard deviations of the first 20 principal components. Genotype imputation was carried out on the Michigan Imputation Server (Das et al., 2016) using Minimac4 and the Haplotype Reference Consortium (HRC) r1.1 2016 (GRCh37/hg19). Post-imputation QC was performed by filtering out variants having a poor imputation quality score (i.e., R^2^<0.3) and MAF <0.05.

### 2.3. Statistical analyses

Polygenic risk scores for BP, MDD, NEU, and SCZ were calculated in each target sample after hard-calling with a genotype probability threshold of 0.9, using PRSice v2.3.3 (https://prsice.info). Summary statistics of the largest genome-wide association studies (GWASs) on BP, MDD, NEU, and SCZ available at the time of conducting our analyses were used as base datasets (Table S1). Clumping was performed to remove SNPs in linkage disequilibrium (r^2^ > 0.1, 250 kb window). Eight *a priori* GWAS P-value thresholds (P_T_) (1e-4, 0.001, 0.05, 0.1, 0.2, 0.3, 0.4, 0.5) were used to select SNPs to be included in each PRS (Choi et al., 2020).

Logistic regressions between each scaled PRS and the two clinical outcomes (non-response and non-remission) were conducted in R v4.0.2, adjusting for age, sex, baseline symptom severity (for non-remission), relevant population principal components, and recruitment sites.

The results obtained in each sample were meta-analysed using the metafor R package (https://cran.r-project.org/web/packages/metafor), within a fixed-effect inverse-variance weighted model, as done by other authors (Zheutlin et al., 2019).

Bonferroni correction was applied considering the four base phenotypes and the eight P_T_used for the PRS calculation and subsequent analyses (α=0.05/(4*8)=1.56e-3).

The PRSs for each of the four base phenotypes showed adequate power (>80%) in predicting both phenotypes in the target samples (Table S1), as assessed through the AVENGEME R package (Palla and Dudbridge, 2015).

## 3. Results

After QC, a total of 3,637 and 3,184 patients were included in the PRS analyses for non-response and non-remission, respectively (non-response: N=2,087; non-remission: N=2,099); details on each target sample are in Table 1.

**Table 1.** Target samples used for the computation of polygenic risk scores and subsequent analyses, after quality control. Abbreviations: N, sample size; Sample prevalence = (N cases/N total); GSRD, European Group for the Study of Resistant Depression; STAR*D, Sequenced Treatment Alternatives to Relieve Depression; SD, standard deviations; *χ*^2^, Pearson’s Chi-squared test statistic with Yates’ continuity correction; t, Welch Two Sample t-test statistic; HAMD_21_, 21-item Hamilton Depression Rating Scale score; MADRS, Montgomery-Åsberg Depression Rating Scale score; QIDS-C_16_, Quick Inventory of Depressive Symptomatology Clinician-rated scale score. * p-value < 0.05.

| Target sample | N controls/cases | Sample prevalence | N total | Age<br>mean (SD)<br>controls/cases | Sex<br>% males<br>controls/cases | Baseline severity<br>mean (SD)<br>controls/cases |
| --- | --- | --- | --- | --- | --- | --- |
| Brescia | response/non-response = 72/381 | 0.84 | 455 | 53.92/56.74<br>(13.27/13.70)<br>t = -1.65, p = 0.10 | 0.26/0.33<br>$\chi^2 = 1.038$, p = 0.31 | MADRS = 30.59/32.01<br>(5.35/5.66)<br>t = -1.86, p = 0.07 |
| GSRD | response/non-response = 279/870 | 0.76 | 1,149 | 51.57/51.93<br>(15.70/13.51)<br>t = -0.34, p = 0.74 | 0.35/0.33<br>$\chi^2 = 0.30$, p = 0.58 | MADRS = 33.16/34.92<br>(7.65/7.54)<br>t = -3.35, p = 8.8e-4* |
| | remission/non-remission = 189/960 | 0.84 | 1,149 | 52.24/51.76<br>(15.48/13.78)<br>t = 0.39, p = 0.69 | 0.32/0.34<br>$\chi^2 = 0.38$, p = 0.54 | MADRS = 32.25/34.94<br>(8.04/7.44)<br>t = -4.24, p = 3.1e-5* |
| Münster | response/non-response = 351/206 | 0.37 | 557 | 49.6/49.7<br>(15.1/16.1)<br>t = -0.03; p = 0.97 | 0.58/0.57<br>$\chi^2 = 1.5e-3$, p = 0.97 | HAMD<sub>21</sub> = 23.06/20.77<br>(7.52/6.73)<br>t = 3.71, p = 2.3e-4* |
| | remission/non-remission = 249/308 | 0.55 | 557 | 49/50.2<br>(15/15.8)<br>t = -0.88, p = 0.38 | 0.57/0.58<br>$\chi^2 = 0$, p = 1 | HAMD<sub>21</sub> = 20.73/23.42<br>(6.94/7.40)<br>t = -4.41, p = 1.3e-5* |
| STAR*D | response/non-response = 795/605 | 0.43 | 1,400 | 42.45/43.78<br>(13.41/13.40)<br>t = -1.89, p = 0.06 | 0.37/0.40<br>$\chi^2 = 1.75$, p = 0.19 | QIDS-C<sub>16</sub> = 16.14/16.52<br>(3.10/3.25)<br>t = -2.23, p = 0.03* |
| | remission/non-remission = 597/803 | 0.57 | 1,400 | 42.23/43.62<br>(13.75/13.14)<br>t = -1.97, p = 0.049* | 0.36/0.40<br>$\chi^2 = 1.85$, p = 0.17 | QIDS-C<sub>16</sub> = 15.62/16.82<br>( 2.97/3.22)<br>t = -7.42, p = 2.0e-13* |
| Tartu | response/non-response = 53/25 | 0.32 | 78 | 30.45/31.92<br>(12.23/10.46)<br>t = -0.55, p = 0.59 | 0.42/0.2<br>$\chi^2 = 2.59$, p = 0.11 | MADRS = 28.17/29.52<br>(5.55/4.61)<br>t = -1.13, p = 0.26 |
| | remission/non-remission = 50/28 | 0.36 | 78 | 29.86/32.82<br>(11.99/10.95)<br>t = -1.11, p = 0.27 | 0.40/0.25<br>$\chi^2 = 1.18$, p = 0.28 | MADRS = 27.38/30.79<br>(4.16/6.34)<br>t = -2.55, p = 0.01* |
| meta-analysis | response/non-response = 1,550/2,087 | 0.57 | 3,637 |  |  |  |
|  | remission/non-remission = 1,085/2,099 | 0.66 | 3,184 |  |  |  |

In the meta-analyses, no association survived Bonferroni correction (Table 2). The top result was found for MDD-PRS, which was nominally positively associated with non-response (p=0.013, pseudo-*R*^2^=0.24%) and non-remission (p=0.004, pseudo-*R*^2^=0.57%) (Table 2 and Table S2-S3). The direction of the association was concordant in three out of the four samples considered (a forest plot is depicted in Fig. 1). Across samples, patients in the highest MDD-PRS quintile showed a higher risk of non-remission than patients in the lowest quintile (OR=1.5, 95% CI 1.11-1.98, p=0.007) (Fig. 2). We found another nominal association between SCZ-PRS and non-remission (p=0.035, pseudo-*R*^2^=0.37%) (Table 2, Table S2 and S6).

**Table 2.**
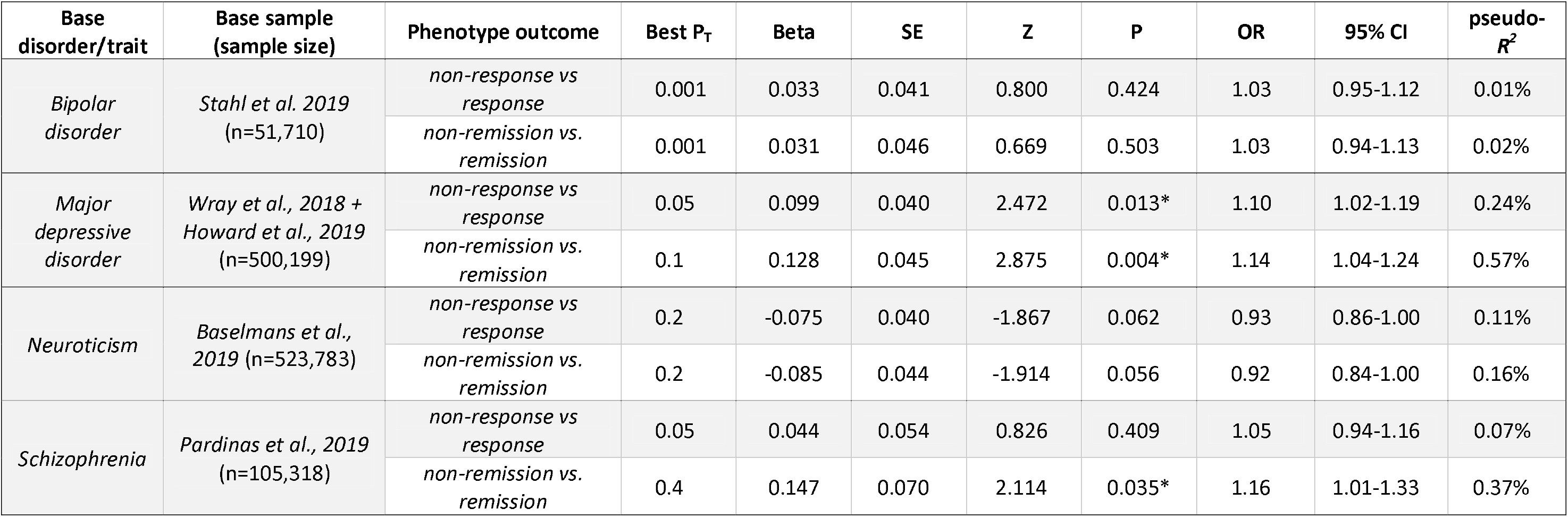
Association between polygenic risk scores (PRSs) and non-response to antidepressants or symptomatologic non-remission in patients with major depressive disorder. Results are shown for the best fitting P-Threshold (achieving the lowest p-value for the association between each PRS and the phenotype outcome). Abbreviations: P_T_, base genome-wide P-Threshold; Beta, estimated coefficient of the model; SE, standard error; Z, test statistics of the coefficient; P, p-value; OR, odds ratio; CI, confidence interval, pseudo-*R*^*2*^, Nagelkerke’s *R*^*2*^. ^*^ Nominally significant results (p-value <0.05). ^**^ Statistically significant result (Bonferroni corrected α = 0.05/(8 genome-wide P-thresholds x 4 base traits/disorders) = 1.56e-3).

**Figure 1.**
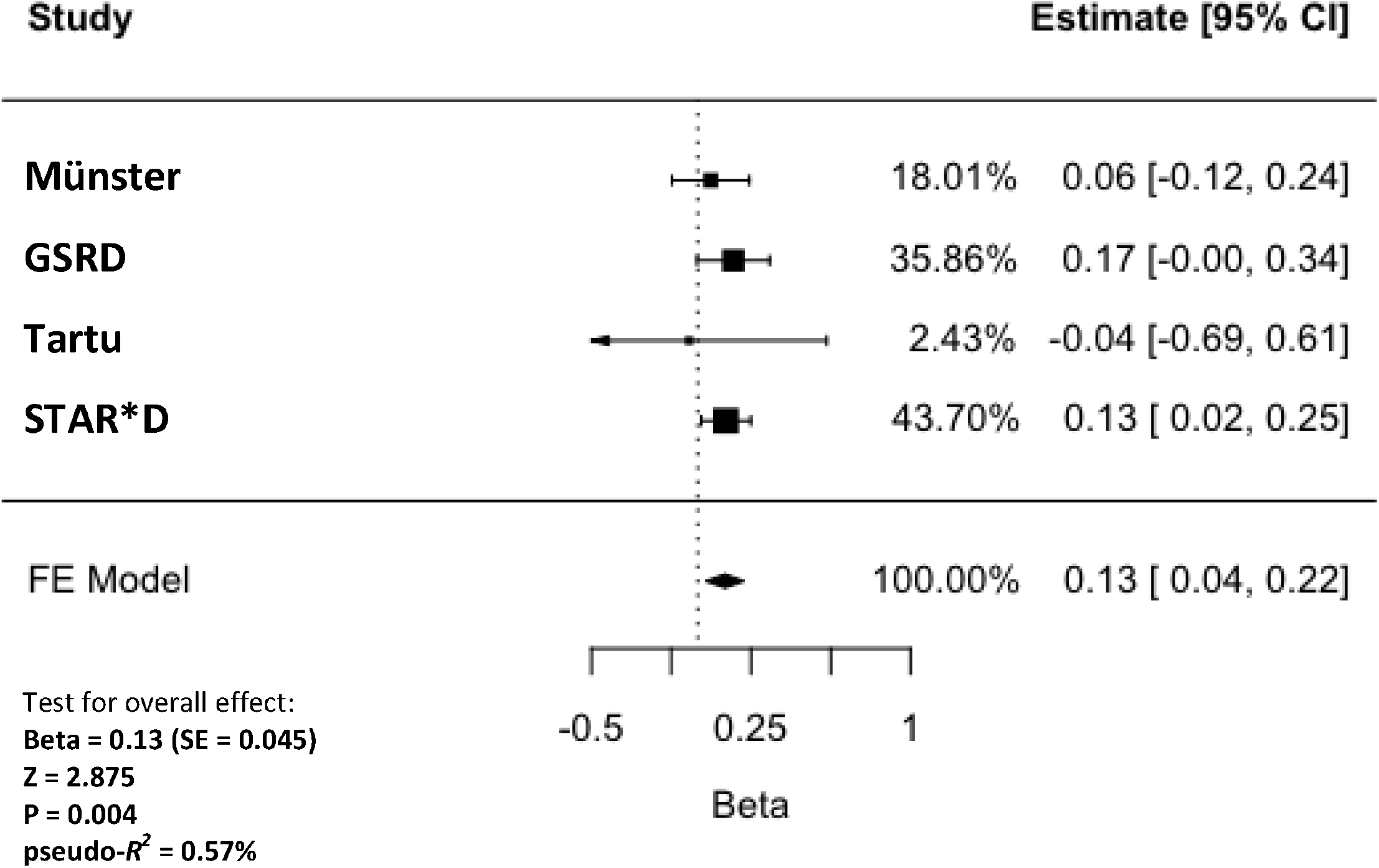
Forest plot showing the association between the polygenic risk score for major depressive disorder (calculated at the genome-wide P-threshold of 0.1) and **non-remission** across the four included clinical cohorts. Abbreviations: FE, fixed effects; Beta, estimated coefficient of the model; SE, standard error; Z, test statistics of the coefficient; P, p-value; OR, odds ratio; CI, confidence interval; pseudo-*R*^*2*^, Nagelkerke’s *R*^*2*^; GSRD, European Group for the Study of Resistant Depression; STAR^*^D, Sequenced Treatment Alternatives to Relieve Depression.

**Figure 2.**
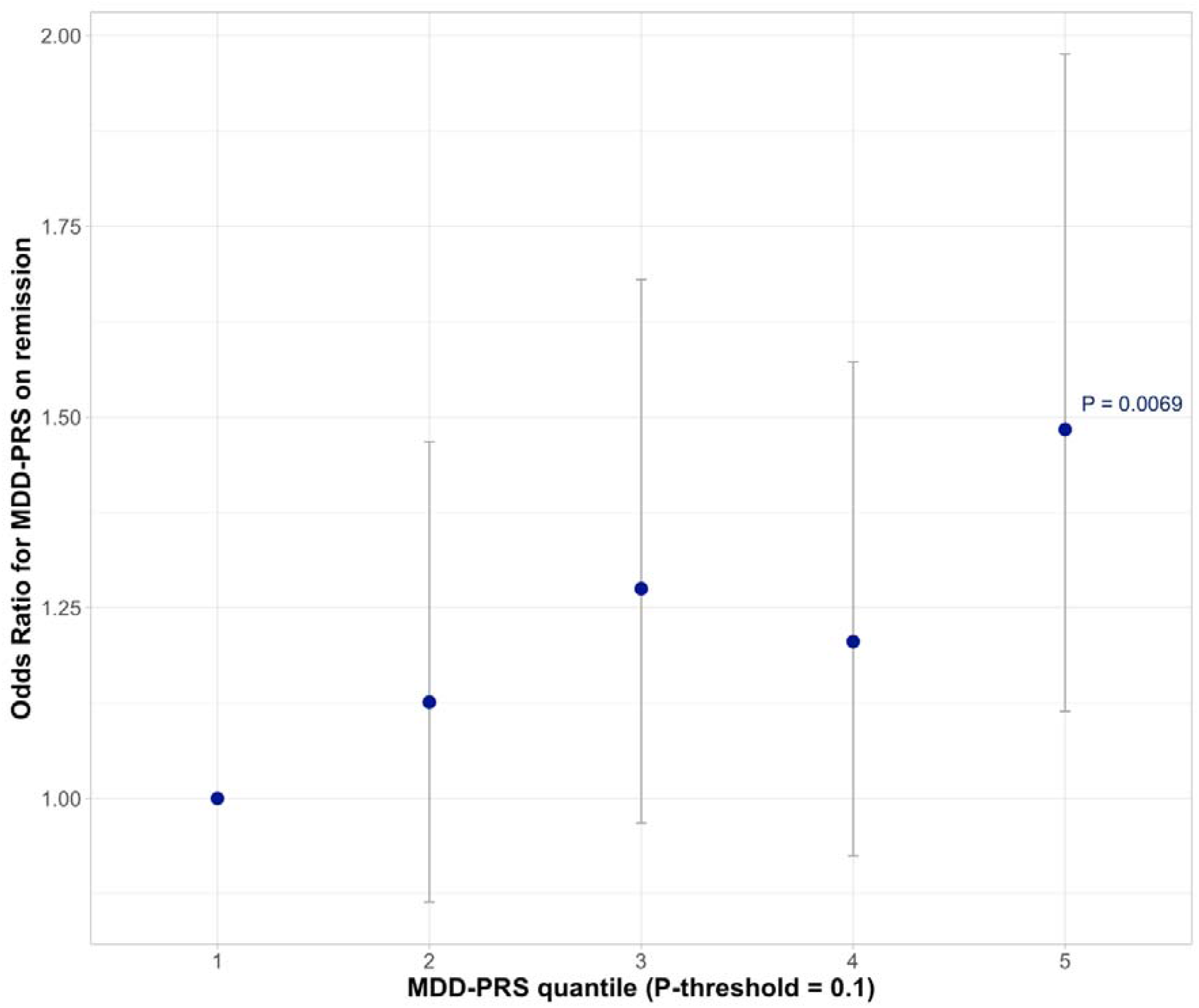
Strata plot showing the odds ratios and related 95% confidence intervals for each MDD-PRS quintile on non-remission, after meta-analysis. The 1^st^ MDD-PRS quintile was used as reference (odds ratio = 1). Abbreviations: MDD-PRS, polygenic risk score for major depressive disorder; P, p-value.

## 4. Discussion

In the present meta-analysis, we investigated whether PRSs for BP, MDD, neuroticism, and SCZ were associated with antidepressant response or remission in a total sample of up to 3,637 patients with MDD. No association survived after Bonferroni correction; nominal associations were found between MDD-PRS, non-remission and non-response, as well as between SCZ-PRS and non-remission.

Though it did not survive a strict multiple-testing correction, a higher genetic burden for MDD may influence remission to antidepressants, an effect that was independent of age, sex, and baseline symptom severity. This result is in line with a previous study in independent samples that examined the degree of treatment resistance as number of failed drug trials (Wigmore et al., 2020). MDD-PRS and PRS for depressive symptoms also showed a trend of association with both symptom improvement and remission in treatment-resistant patients treated with esketamine addon (Li et al., 2020). Similarly, MDD-PRS may impact the risk of treatment with electroconvulsive therapy, an option of choice in TRD (Foo et al., 2019). On the other hand, a previous study showing overlap with the present target sample did not identify any association between MDD-PRS and symptom improvement to antidepressants (Garcia-Gonzalez et al., 2017); no significant genetic covariance between MDD and antidepressant treatment outcomes was found by another study (Pain et al., 2020).

The nominal association between higher SCZ-PRS and non-remission is in line with a previous study indicating negative genetic covariance between SCZ and remission (Pain et al., 2020). Although our meta-analysis did not find an association between SCZ-PRS and non-response, the analysis limited to the GSRD sample had previously suggested a negative effect of polygenic risk for SCZ on antidepressant response (Fanelli et al., 2021). This difference may be explained by the higher baseline symptom severity of patients in the GSRD sample compared to patients in the other samples included in the present meta-analysis. Indeed, previous evidence indicates that higher SCZ-PRSs are associated with higher severity of symptoms and functional impairment in MDD (Fanelli et al., 2021). Of note, the response phenotype considered in the present meta-analysis was defined based on the last antidepressant treatment, while in our previous study we considered separately non-responders to the last treatment and TRD (non-responders to two or more treatments). This study comes with some strengths and limitations. Noteworthy is the use of a standardised genetic quality control strategy on the target datasets in line with current Psychiatric Genomics Consortium standards, the use of a strict correction for multiple testing to minimise type 1 errors, as well as the inclusion of socio-demographic and clinical variables such as age, sex and depressive symptoms severity at baseline as potential confounders in the regression models. Nevertheless, our results could be subject to potential overfitting, and an out-of-sample validation was not performed, as we preferred a meta-analytic approach to optimise power. A limitation may have arisen from having considered an early time point of six weeks for the assessment of remission in some of the samples included in the meta-analysis, possibly underestimating remission rates.

In conclusion, although PRSs are not able to significantly predict treatment response or remission in MDD yet, our study suggests an increased genetic susceptibility to MDD and SCZ in patients who do not achieve remission/response after the first antidepressant treatment, in line with the previous literature. Integration of PRSs with clinical predictors may in the future provide an opportunity for better prediction of outcomes and optimisation of treatment choices.

## Data Availability

Access to the Sequenced Treatment Alternatives to Relieve Depression (STAR*D) data is granted to Principal Investigators after submission of a research proposal to the National Institute of Mental Health (NIMH) via the NIMH Repository & Genomics Resource (NRGR) (https://www.nimhgenetics.org).

## Statement of Ethics

The authors assert that all procedures contributing to this work comply with the ethical standards of the relevant national and institutional committees on human experimentation and with the Helsinki Declaration of 1975, as revised in 2008. All subjects selected by clinicians were included in the screening phase after obtaining their written informed consent. In more detail, the Brescia study protocol was approved by the Ethics Committee of the province of Verona, Italy; the European Group for the Study of Resistant Depression (GRSD) study protocol was approved by the Ethics Committee of the coordinating centre at Hôpital Erasme, Cliniques universitaires de Bruxelles (Université Libre de Bruxelles), Belgium, and the local ethical committees of all the other nine participating European academic centres; the Münster study protocol was approved by the ethical board of the University of Münster, Germany; the Tartu study protocol was approved by the Human Studies Ethics Committee of the University of Tartu and State Agency of Medicines; the Sequenced Treatment Alternatives to Relieve Depression (STAR*D) study protocol received ethics approval from 14 participating institutional review boards, a National Coordinating Center, a Data Coordinating Center, and the Data Safety and Monitoring Board at the National Institute of Mental Health (NIMH), Bethesda, Maryland, US. This research group certifies that data collected for the STAR*D were exclusively used for scientific investigation. Before obtaining access to data, the objectives of our investigation were clearly described in the request form.

## Contributors

Giuseppe Fanelli contributed to the conceptualisation of the study, performed the analyses, interpreted the results and wrote the first draft of the manuscript. Alessandro Serretti and Chiara Fabbri conceptualised the study, helped with the interpretation of the results, reviewed the first draft of the manuscript and contributed to the funding acquisition. Chiara Fabbri supervised the whole process leading to the final publication. The other authors contributed to data collection, data preparation and/or the improvement of the final version of the paper. All named authors meet the International Committee of Medical Journal Editors (ICMJE) criteria for authorship for this manuscript, take responsibility for the integrity of the work as a whole, and have given final approval for the version to be published.

## Acknowledgments

The European College of Neuropsychopharmacology (ECNP) Pharmacogenomics & Transcriptomics Thematic Working Group commissioned this manuscript and contributed by sharing individual genotyped data, as well as providing comments and critical review to the manuscript.

We thank the US National Institute of Mental Health (NIMH) for providing access to data on the Sequenced Treatment Alternatives to Relieve Depression (STAR*D) sample. We also thank the authors of previous publications in this dataset, and foremost, we thank the patients and their families who agreed to be enrolled in the study. Data were obtained from the limited access dataset distributed from the NIH-supported STAR*D (ClinicalTrials.gov ID NCT00021528; Request ID 5ce26a95712d8). The study was supported by NIMH Contract No. N01MH90003 to the University of Texas Southwestern Medical Center.

We thank the European Group for the Study of Resistant Depression (GRSD) for providing access to the dataset they collected, as well as the researchers of the consortia that provided the GWAS summary statistics used in our analyses and the participants of the cohorts to which they refer.

## References

Aluoja, A., Toru, I., Raag, M., Eller, T., Vohma, U., Maron, E., 2018. Personality traits and escitalopram treatment outcome in major depression. Nord J Psychiatry 72, 354–360.

Baune, B.T., Dannlowski, U., Domschke, K., Janssen, D.G., Jordan, M.A., Ohrmann, P., Bauer, J., Biros, E., Arolt, V., Kugel, H., Baxter, A.G., Suslow, T., 2010. The interleukin 1 beta (IL1B) gene is associated with failure to achieve remission and impaired emotion processing in major depression. Biol Psychiatry 67, 543–549.

Choi, S.W., Mak, T.S., O’Reilly, P.F., 2020. Tutorial: a guide to performing polygenic risk score analyses. Nat Protoc 15, 2759–2772.

Das, S., Forer, L., Schonherr, S., Sidore, C., Locke, A.E., Kwong, A., Vrieze, S.I., Chew, E.Y., Levy, S., McGue, M., Schlessinger, D., Stambolian, D., Loh, P.R., Iacono, W.G., Swaroop, A., Scott, L.J., Cucca, F., Kronenberg, F., Boehnke, M., Abecasis, G.R., Fuchsberger, C., 2016. Next-generation genotype imputation service and methods. Nat Genet 48, 1284–1287.

De Carlo, V., Calati, R., Serretti, A., 2016. Socio-demographic and clinical predictors of non-response/non-remission in treatment resistant depressed patients: A systematic review. Psychiatry Res 240, 421–430.

Diseases, G.B.D., Injuries, C., 2020. Global burden of 369 diseases and injuries in 204 countries and territories, 1990-2019: a systematic analysis for the Global Burden of Disease Study 2019. Lancet 396, 1204–1222.

Fanelli, G., Benedetti, F., Kasper, S., Zohar, J., Souery, D., Montgomery, S., Albani, D., Forloni, G., Ferentinos, P., Rujescu, D., Mendlewicz, J., Serretti, A., Fabbri, C., 2021. Higher polygenic risk scores for schizophrenia may be suggestive of treatment non-response in major depressive disorder. Prog Neuropsychopharmacol Biol Psychiatry 108, 110170.

Foo, J.C., Streit, F., Frank, J., Witt, S.H., Treutlein, J., Major Depressive Disorder Working Group of the Psychiatric Genomics, C., Baune, B.T., Moebus, S., Jockel, K.H., Forstner, A.J., Nothen, M.M., Rietschel, M., Sartorius, A., Kranaster, L., 2019. Evidence for increased genetic risk load for major depression in patients assigned to electroconvulsive therapy. Am J Med Genet B Neuropsychiatr Genet 180, 35–45.

Garcia-Gonzalez, J., Tansey, K.E., Hauser, J., Henigsberg, N., Maier, W., Mors, O., Placentino, A., Rietschel, M., Souery, D., Zagar, T., Czerski, P.M., Jerman, B., Buttenschon, H.N., Schulze, T.G., Zobel, A., Farmer, A., Aitchison, K.J., Craig, I., McGuffin, P., Giupponi, M., Perroud, N., Bondolfi, G., Evans, D., O’Donovan, M., Peters, T.J., Wendland, J.R., Lewis, G., Kapur, S., Perlis, R., Arolt, V., Domschke, K., Major Depressive Disorder Working Group of the Psychiatric Genomic, C., Breen, G., Curtis, C., Sang-Hyuk, L., Kan, C., Newhouse, S., Patel, H., Baune, B.T., Uher, R., Lewis, C.M., Fabbri, C., 2017. Pharmacogenetics of antidepressant response: A polygenic approach. Prog Neuropsychopharmacol Biol Psychiatry 75, 128–134.

Howland, R.H., 2008. Sequenced Treatment Alternatives to Relieve Depression (STAR*D). Part 1: study design. J Psychosoc Nurs Ment Health Serv 46, 21–24.

Kautzky, A., Dold, M., Bartova, L., Spies, M., Vanicek, T., Souery, D., Montgomery, S., Mendlewicz, J., Zohar, J., Fabbri, C., Serretti, A., Lanzenberger, R., Kasper, S., 2018. Refining Prediction in Treatment-Resistant Depression: Results of Machine Learning Analyses in the TRD III Sample. J Clin Psychiatry 79.

Lam, M., Awasthi, S., Watson, H.J., Goldstein, J., Panagiotaropoulou, G., Trubetskoy, V., Karlsson, R., Frei, O., Fan, C.C., De Witte, W., Mota, N.R., Mullins, N., Brugger, K., Lee, S.H., Wray, N.R., Skarabis, N., Huang, H., Neale, B., Daly, M.J., Mattheisen, M., Walters, R., Ripke, S., 2020. RICOPILI: Rapid Imputation for COnsortias PIpeLIne. Bioinformatics 36, 930–933.

Li, Q.S., Wajs, E., Ochs-Ross, R., Singh, J., Drevets, W.C., 2020. Genome-wide association study and polygenic risk score analysis of esketamine treatment response. Sci Rep 10, 12649.

Minelli, A., Magri, C., Barbon, A., Bonvicini, C., Segala, M., Congiu, C., Bignotti, S., Milanesi, E., Trabucchi, L., Cattane, N., Bortolomasi, M., Gennarelli, M., 2015. Proteasome system dysregulation and treatment resistance mechanisms in major depressive disorder. Transl Psychiatry 5, e687.

Natarajan, P., Young, R., Stitziel, N.O., Padmanabhan, S., Baber, U., Mehran, R., Sartori, S., Fuster, V., Reilly, D.F., Butterworth, A., Rader, D.J., Ford, I., Sattar, N., Kathiresan, S., 2017. Polygenic Risk Score Identifies Subgroup With Higher Burden of Atherosclerosis and Greater Relative Benefit From Statin Therapy in the Primary Prevention Setting. Circulation 135, 2091–2101.

Pain, O., Hodgson, K., Trubetskoy, V., Ripke, S., Marshe, V.S., Adams, M.J., Byrne, E.M., Campos, A.I., Carrillo-Roa, T., Cattaneo, A., Als, T.D., Souery, D., Dernovsek, M.Z., Fabbri, C., Hayward, C., Henigsberg, N., Hauser, J., Kennedy, J.L., Lenze, E.J., Lewis, G., Müller, D.J., Martin, N.G., Mulsant, B.H., Mors, O., Perroud, N., Porteous, D.J., RenterÍa, M.E., Reynolds, C.F., Rietschel, M., Uher, R., Wigmore, E.M., Maier, W., Wray, N.R., Aitchison, K.J., Arolt, V., Baune, B.T., Biernacka, J.M., Bondolfi, G., Domschke, K., Kato, M., Li, Q.S., Liu, Y.-L., Serretti, A., Tsai, S.-J., Turecki, G., Weinshilboum, R., McIntosh, A.M., Lewis, C.M., 2020. Antidepressant Response in Major Depressive Disorder: A Genome-wide Association Study. medRxiv, 2020.2012.2011.20245035.

Palla, L., Dudbridge, F., 2015. A Fast Method that Uses Polygenic Scores to Estimate the Variance Explained by Genome-wide Marker Panels and the Proportion of Variants Affecting a Trait. Am J Hum Genet 97, 250–259.

Rush, A.J., Trivedi, M.H., Wisniewski, S.R., Nierenberg, A.A., Stewart, J.W., Warden, D., Niederehe, G., Thase, M.E., Lavori, P.W., Lebowitz, B.D., McGrath, P.J., Rosenbaum, J.F., Sackeim, H.A., Kupfer, D.J., Luther, J., Fava, M., 2006. Acute and longer-term outcomes in depressed outpatients requiring one or several treatment steps: a STAR*D report. Am J Psychiatry 163, 1905–1917.

Souery, D., Oswald, P., Massat, I., Bailer, U., Bollen, J., Demyttenaere, K., Kasper, S., Lecrubier, Y., Montgomery, S., Serretti, A., Zohar, J., Mendlewicz, J., Group for the Study of Resistant, D., 2007. Clinical factors associated with treatment resistance in major depressive disorder: results from a European multicenter study. J Clin Psychiatry 68, 1062–1070.

Tammiste, A., Jiang, T., Fischer, K., Magi, R., Krjutskov, K., Pettai, K., Esko, T., Li, Y., Tansey, K.E., Carroll, L.S., Uher, R., McGuffin, P., Vosa, U., Tsernikova, N., Saria, A., Ng, P.C., Eller, T., Vasar, V., Nutt, D.J., Maron, E., Wang, J., Metspalu, A., 2013. Whole-exome sequencing identifies a polymorphism in the BMP5 gene associated with SSRI treatment response in major depression. J Psychopharmacol 27, 915–920.

Wigmore, E.M., Hafferty, J.D., Hall, L.S., Howard, D.M., Clarke, T.K., Fabbri, C., Lewis, C.M., Uher, R., Navrady, L.B., Adams, M.J., Zeng, Y., Campbell, A., Gibson, J., Thomson, P.A., Hayward, C., Smith, B.H., Hocking, L.J., Padmanabhan, S., Deary, I.J., Porteous, D.J., Mors, O., Mattheisen, M., Nicodemus, K.K., McIntosh, A.M., 2020. Genome-wide association study of antidepressant treatment resistance in a population-based cohort using health service prescription data and meta-analysis with GENDEP. Pharmacogenomics J 20, 329–341.

Zheutlin, A.B., Dennis, J., Karlsson Linner, R., Moscati, A., Restrepo, N., Straub, P., Ruderfer, D., Castro, V.M., Chen, C.Y., Ge, T., Huckins, L.M., Charney, A., Kirchner, H.L., Stahl, E.A., Chabris, C.F., Davis, L.K., Smoller, J.W., 2019. Penetrance and Pleiotropy of Polygenic Risk Scores for Schizophrenia in 106,160 Patients Across Four Health Care Systems. Am J Psychiatry 176, 846–855.

